# Early assessment of the clinical severity of the SARS-CoV-2 Omicron variant in South Africa

**DOI:** 10.1101/2021.12.21.21268116

**Authors:** Nicole Wolter, Waasila Jassat, Sibongile Walaza, Richard Welch, Harry Moultrie, Michelle Groome, Daniel Gyamfi Amoako, Josie Everatt, Jinal N. Bhiman, Cathrine Scheepers, Naume Tebeila, Nicola Chiwandire, Mignon du Plessis, Nevashan Govender, Arshad Ismail, Allison Glass, Koleka Mlisana, Wendy Stevens, Florette K. Treurnicht, Zinhle Makatini, Nei-yuan Hsiao, Raveen Parboosing, Jeannette Wadula, Hannah Hussey, Mary-Ann Davies, Andrew Boulle, Anne von Gottberg, Cheryl Cohen

**Author notes:** These authors contributed equally (Anne von Gottberg, Cheryl Cohen). **Corresponding author:** Dr Nicole Wolter Centre for Respiratory Diseases and Meningitis, National Institute for Communicable Diseases, of the National Health Laboratory Service Private Bag X4, Sandringham, 2131, Gauteng, South Africa. **Alternative corresponding author:** Prof Cheryl Cohen Centre for Respiratory Diseases and Meningitis, National Institute for Communicable Diseases, of the National Health Laboratory Service Private Bag X4, Sandringham, 2131, Gauteng, South Africa.

## Abstract

**Background:** The SARS-CoV-2 Omicron variant of concern (VOC) almost completely replaced other variants in South Africa during November 2021, and was associated with a rapid increase in COVID-19 cases. We aimed to assess clinical severity of individuals infected with Omicron, using S Gene Target Failure (SGTF) on the Thermo Fisher Scientific TaqPath COVID-19 PCR test as a proxy.

**Methods:** We performed data linkages for (i) SARS-CoV-2 laboratory tests, (ii) COVID-19 case data, (iii) genome data, and (iv) the DATCOV national hospital surveillance system for the whole of South Africa. For cases identified using Thermo Fisher TaqPath COVID-19 PCR, infections were designated as SGTF or non-SGTF. Disease severity was assessed using multivariable logistic regression models comparing SGTF-infected individuals diagnosed between 1 October to 30 November to (i) non-SGTF in the same period, and (ii) Delta infections diagnosed between April and November 2021.

**Results:** From 1 October through 6 December 2021, 161,328 COVID-19 cases were reported nationally; 38,282 were tested using TaqPath PCR and 29,721 SGTF infections were identified. The proportion of SGTF infections increased from 3% in early October (week 39) to 98% in early December (week 48). On multivariable analysis, after controlling for factors associated with hospitalisation, individuals with SGTF infection had lower odds of being admitted to hospital compared to non-SGTF infections (adjusted odds ratio (aOR) 0.2, 95% confidence interval (CI) 0.1-0.3). Among hospitalised individuals, after controlling for factors associated with severe disease, the odds of severe disease did not differ between SGTF-infected individuals compared to non-SGTF individuals diagnosed during the same time period (aOR 0.7, 95% CI 0.3-1.4). Compared to earlier Delta infections, after controlling for factors associated with severe disease, SGTF-infected individuals had a lower odds of severe disease (aOR 0.3, 95% CI 0.2-0.5).

**Conclusion:** Early analyses suggest a reduced risk of hospitalisation among SGTF-infected individuals when compared to non-SGTF infected individuals in the same time period. Once hospitalised, risk of severe disease was similar for SGTF- and non-SGTF infected individuals, while SGTF-infected individuals had a reduced risk of severe disease when compared to earlier Delta-infected individuals. Some of this reducton is likely a result of high population immunity.

## INTRODUCTION

Since the introduction of severe acute respiratory syndrome coronavirus-2 (SARS-CoV-2) in March 2020, South Africa has experienced three epidemic waves with the Beta and Delta variants of concern (VOCs) dominating the second and third waves, respectively. On 24 November 2021, the Network for Genomics Surveillance of South Africa (NGS-SA) reported a new variant of SARS-CoV-2 which had been detected from specimens collected on 14 November 2021 in South Africa, originally assigned to the lineage B.1.1.529, to the World Health Organization (WHO)^1^. The WHO, on the recommendation of the Technical Advisory Group on SARS-CoV-2 Virus Evolution, designated B.1.1.529 as Omicron^2^, the fifth VOC, on 26^th^ November 2021. Concomitantly, there was a rapid increase in COVID-19 cases in Gauteng Province^1^, from the week beginning 15 November 2021, with SARS-CoV-2 testing laboratories reporting an increase in the number of samples with S gene target failure (SGTF) when tested on the TaqPath™ COVID-19 (Thermo Fisher Scientific, Waltham, MA, USA) PCR test. Subsequently, increases in COVID-19 cases and samples with SGTF were observed in other provinces in South Africa precipitating entry into a fourth wave of SARS-CoV-2 infections. By 16 December 2021, Omicron had been detected in 87 countries, with many countries reporting community transmission.

The Omicron VOC has a high number of mutations, some of which are concerning due to predicted immune evasion and increased infectivity. While the Omicron variant shares a few common mutations with the C.1.2 (a highly mutated lineage previously identified in South Africa)^3^, Beta and Delta variants, it also has 22 additional substitutions (including insertions and deletions) not seen in any other VOC or variant of interest (VOI) to date. Among them, the Δ69-70 amino acid deletion in the spike gene, previously observed in Alpha, is known to cause SGTF on the TaqPath COVID-19 PCR test.

Data on the clinical severity of the Omicron variant compared to previous SARS-CoV-2 variants are needed to guide public health planning and response. DATCOV-Gen^4^ is a prospective surveillance network linking real-time SARS-CoV-2 genome data to detailed epidemiologic and clinical data on hospitalised cases to allow rapid assessment of severity and clinical presentation of emerging SARS-CoV-2 VOCs. We aimed to assess the severity of Omicron infections compared to Delta variant using SGTF as a proxy^5^.

## METHODS

### Data source and linkage

We linked data from four sources: (i) National COVID-19 case data reported in real-time to the National Institute for Communicable Diseases (NICD) Notifiable Medical Conditions Surveillance System (NMCSS), (ii) SARS-CoV-2 laboratory test data (test used and PCR cycle threshold (Ct) values) for the period 1 October – 6 December 2021 for public sector laboratories (National Health Laboratory Service (NHLS)) reported all test data) and one large private sector laboratory (reported TaqPath PCR test only), (iii) genome data for clinical specimens sent to NICD from private and public diagnostic laboratories around the country (predominantly from Gauteng, North West, Mpumalanga and Northern Cape provinces), and collected through the pneumonia surveillance programme^6^ in five provinces (Western Cape, KwaZulu-Natal, North West, Gauteng and Mpumalanga), and (iv) DATCOV, which is an active surveillance system for COVID-19 hospital admissions with comprehensive coverage of all hospitals in South Africa^7^. DATCOV includes patients with COVID-19 symptoms, acquired nosocomial COVID-19 infection, or tested positive incidentally when admitted for other reasons. Case and test data were obtained on 6 December 2021, and DATCOV data on 21 December 2021. For hospitalisation and severity analyses, cases were censored to those with a specimen collected before 1 December 2021.

### Definitions

Infections were classified as SGTF (as a proxy for Omicron) when an individual tested positive using the TaqPath COVID-19 PCR test with non-detectable S gene target and Ct value ≤30 for either the ORF1ab or nucleocapsid (N) gene targets^8^. Infections were classified as non-SGTF when an individual tested positive using TaqPath COVID-19 PCR test with Ct≤30 for either the ORF1ab or nucleocapsid (N) gene targets and had detectable S gene target. An individual was classified as admitted to hospital if they linked to a case on the DATCOV database with an admission date from 7 days prior to 21 days following the date of specimen collection. Severe disease was defined as a hospitalised patient meeting at least one of the following criteria: admitted to the intensive care unit (ICU), received oxygen treatment, ventilated, received extracorporeal membrane oxygenation (ECMO), experienced acute respiratory distress syndrome (ARDS) and/or died. Co-morbidity was defined as ≥1 of the following conditions: hypertension, diabetes, chronic cardiac disease, chronic kidney disease, asthma, chronic obstructive pulmonary disease (COPD), malignancy, HIV, and active or past tuberculosis. The Delta variant was identified by genome sequencing. Re-infection was defined as an individual with at least one positive test >90 days prior to the current episode.

### Statistical analysis

Analysis was performed using Stata 14.1^®^ (StataCorp LP, College Station, US). Categorical variables were summarised using frequency distributions and compared using Pearson’s Chi-squared test.

The mean Ct value (used as a proxy for viral load) for all public sector positive PCR tests (any PCR test) during the early Omicron wave period (weeks 46-48, 14 November – 4 December 2021) was compared to the early Delta wave period (weeks 18-20, 2-22 May 2021) using the students t-test. The early wave periods were defined from the week before the country crossed a weekly incidence risk of 30 cases per 100,000 persons until two weeks later^9^. Where the PCR test included ≥1 gene target, the target with the lowest Ct value was used for the analysis.

Severity of Omicron was assessed in two ways: (i) by comparing SGTF and non-SGTF infections diagnosed between 1 October to 30 November 2021, and (ii) by comparing Delta variant infections diagnosed during April through November 2021, to SGTF infections that were diagnosed during 1 October to 30 November 2021. Hospitalisation and severity data were obtained for cases on 21 December 2021 to allow at least three weeks follow up period for admission to hospital and in-hospital outcome. Severity analyses were restricted to admissions that had already accumulated outcomes and all patients still in-hospital were excluded, because they were at risk of still developing severe outcomes or death. Among DATCOV cases hospitalised between 5 March 2020-18 December 2021, the median time from admission to in-hospital outcome was 6 days (interquartile range (IQR) 3-11 days, n=414,149); median of 5 days (IQR 2-11 days, n=94,938) for individuals that died and median of 6 days (IQR 3-10, n=304,845) for individuals that were discharged alive.

### (i) SGTF vs. non-SGTF analysis

Two models were generated to assess (i) hospitalisation and (ii) severe disease among hospitalised individuals as outcome variables. Multivariable logistic regression was performed to evaluate the association of SGTF infection, compared to non-SGTF infection, with hospitalisation among cases diagnosed between 1 October – 30 November 2021. We controlled for factors known to be associated with hospitalisation (age, sex, presence of co-morbidity, province and healthcare sector) and adjusted for known prior SARS-CoV-2 infection. Multivariable logistic regression was performed to evaluate the association of SGTF infection, compared to non-SGTF infection, with disease severity among hospitalised individuals with a diagnosis between 1 October – 30 November 2021, restricted to individuals with a known in-hospital outcome on 21 December 2021 (excluding cases still in hospital). We controlled for factors known to be associated with severity (age, presence of co-morbidity, sex, province and healthcare sector) and adjusted for the number of days between the date of specimen collection and date of hospital admission, known prior SARS-CoV-2 infection and SARS-CoV-2 vaccination status.

### (ii) Delta vs. SGTF analysis

For these analyses, we compared Delta variant infections diagnosed between April – November 2021 to SGTF infections diagnosed between 1 October – 30 November 2021. Multivariable logistic regression was performed to evaluate the association of SGTF infection, compared to Delta variant infection, with disease severity among hospitalised individuals, restricted to individuals with a known in-hospital outcome on 21 December 2021 (excluding cases still in hospital). We controlled for factors known to be associated with disease severity (age, presence of co-morbidity, sex, province and healthcare sector), and adjusted for number of days between date of specimen collection and date of hospital admission, known prior SARS-CoV-2 infection and SARS-CoV-2 vaccination status.

### Ethical approval

Ethical approval was obtained from the Human Research Ethics Committee (Medical) of University of the Witwatersrand for the collection of COVID-19 case and test data as part of essential communicable disease surveillance (M160667), and for the DATCOV surveillance programme (M2010108).

## RESULTS

From 1 October (week 39) to 6 December (week 48) 2021, 161,328 COVID-19 cases were reported nationally; 67,988 (42%) from the public sector and 93,340 (58%) from the private sector. Of these, 120,110 (74%) cases were detected by PCR; 38,282 (32%) of which were known to have been tested with the TaqPath PCR test. Among tests performed using TaqPath PCR, 31,133 (81%) had a Ct value ≤30 for at least one target; 29,721 (95%) were SGTF infections and 1412 (5%) were non-SGTF infections. All SGTF samples with genome data as of 14 December 2021 were confirmed as Omicron (n=30)). The proportion of SGTF infections increased from 3% in week 39 to 98% in week 48 of 2021. The proportion of SGTF infections started to increase first in Gauteng province in week 44 (week starting 21 October), followed by Limpopo Province in week 45 (Figure 1). Sharp increases in the proportion of SGTF infections were subsequently detected in all other provinces, and by week 47, the majority of infections in all provinces were SGTF.

**Figure 1.**
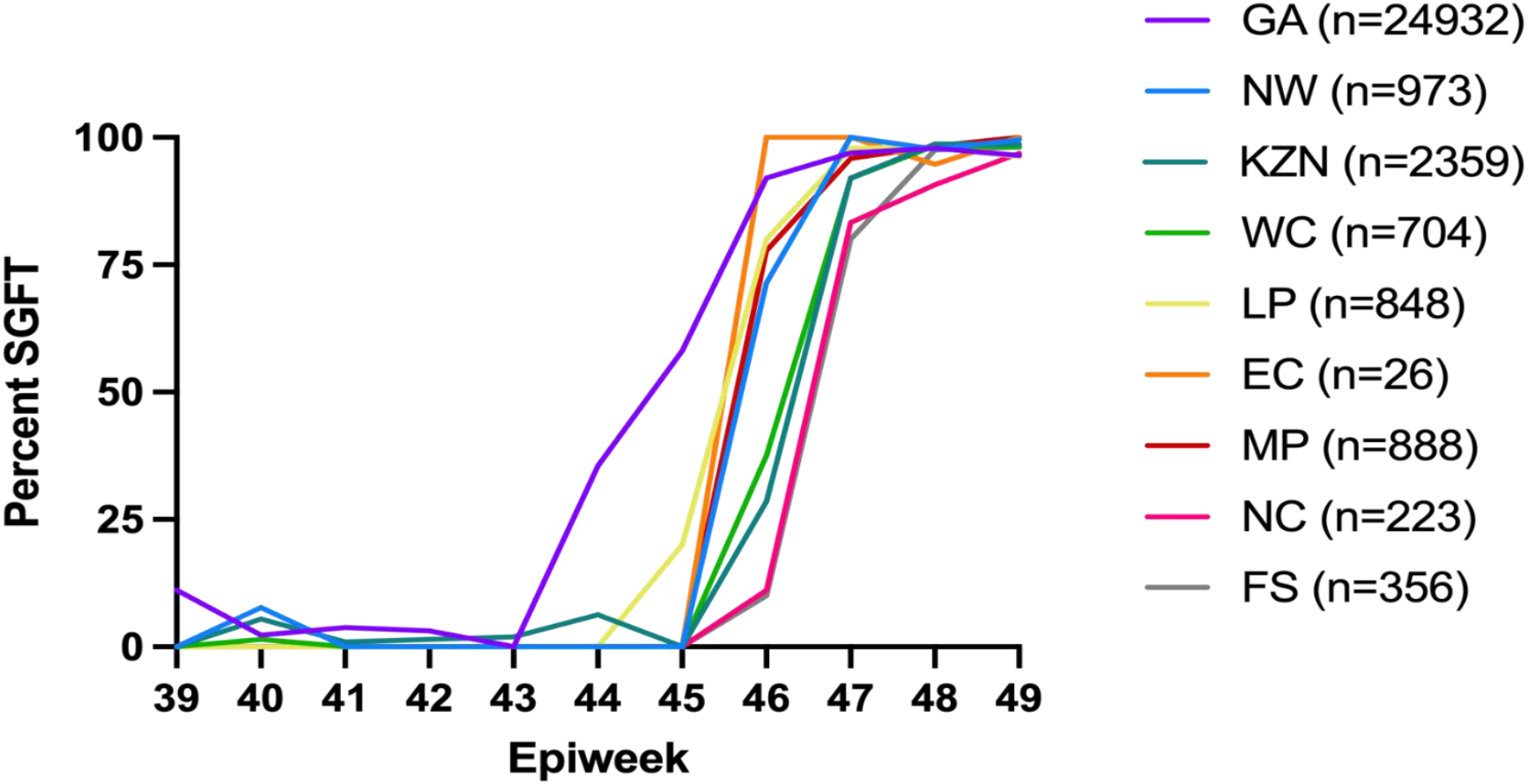
Percentage of S gene target failure (SGTF) infections among tests performed on the TaqPath assay by epidemiological week and province, DATCOV-Gen, 1 October – 6 December 2021. GA, Gauteng; NW, North West; KZN, KwaZulu-Natal; WC, Western Cape; LP, Limpopo; EC, Eastern Cape; MP, Mpumalanga; NC, Northern Cape; FS, Free State

The mean Ct value (± standard deviation) for all positive PCR tests in the public sector in the early Omicron wave period (23.95±6.06, n=16,542) was lower than in the early Delta wave period (26.98±6.52, n=10,022) (P<0.001).

### SGTF vs. non-SGTF infection analysis

Among individuals with SGTF infection diagnosed between 1 October - 30 November 2021, 2.5% (261/10,547) were admitted to hospital, compared to 12.8% (121/948) of individuals with non-SGTF infection (P<0.001). On multivariable analysis, after controlling for factors associated with hospitalisation, individuals with SGTF infection had lower odds of being admitted to hospital compared to non-SGTF infections (adjusted odds ratio (aOR) 0.2, 95% confidence interval (CI) 0.1-0.3) (Table 1). In addition to geographic factors, hospital admission was associated with young age (<5 years, aOR 9.3, 95% CI 5.2-16.8) and older age (≥60 years, aOR 3.1, 95% CI 1.9-5.0) compared to individuals aged 19-24 years and female sex (aOR 1.3, 95% CI 1.1-1.6), whereas individuals in the private healthcare sector were less likely to be admitted to hospital (aOR 0.8, 95% CI 0.6-1.0) compared to the public sector.

**Table 1.**
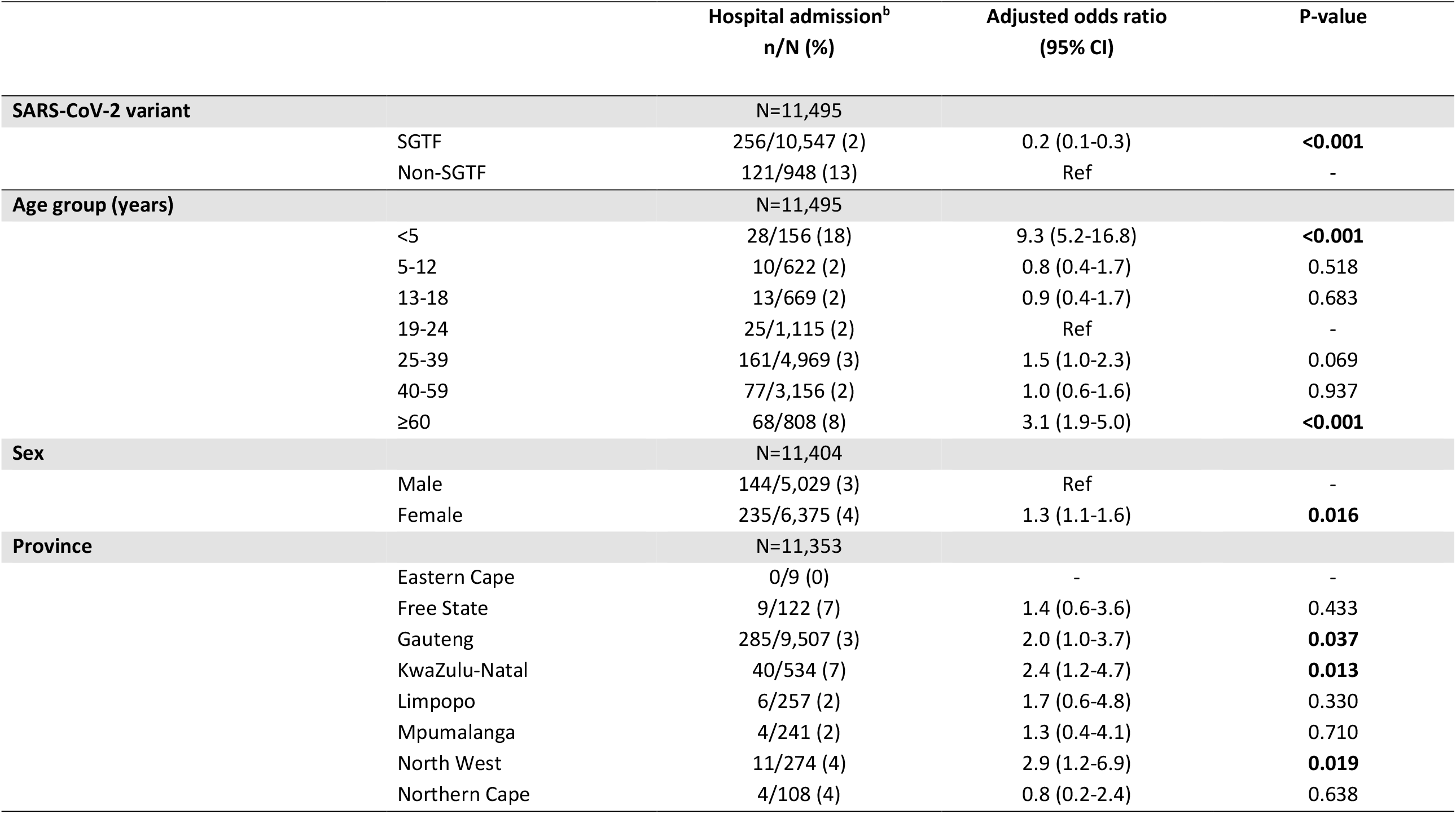
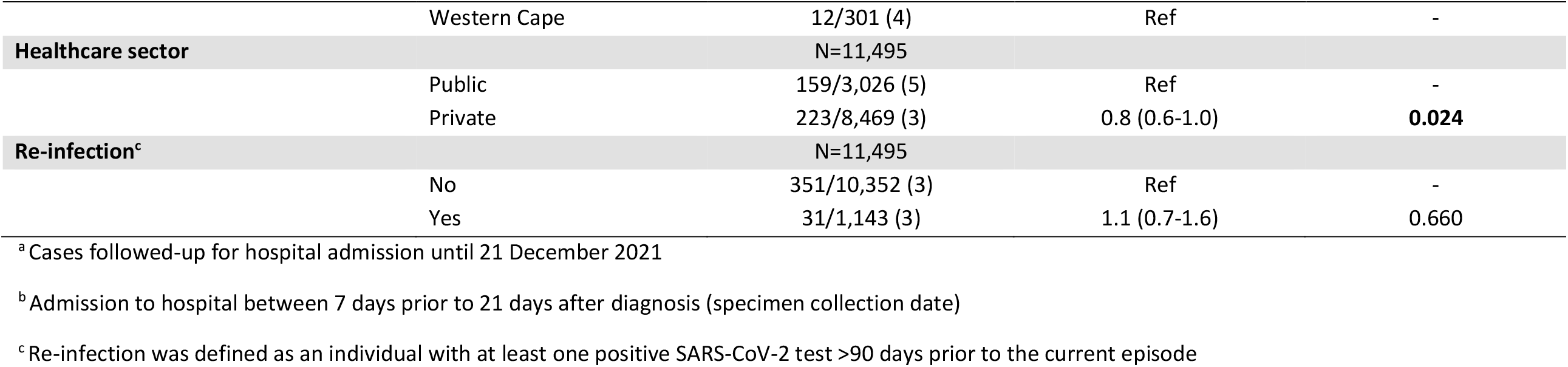
Multivariable logistic regression analysis evaluating the association between S gene target failure (SGTF) infection, compared to non-SGTF infection, and hospitalisation, South Africa, 1 October – 30 November 2021^a^ (N=11,255)

|  | Hospital admission<sup>b</sup><br>n/N (%) | Adjusted odds ratio<br>(95% CI) | P-value |
| --- | --- | --- | --- |
| **SARS-CoV-2 variant** | N=11,495 |  |  |
| SGTF | 256/10,547 (2) | 0.2 (0.1-0.3) | **<0.001** |
| Non-SGTF | 121/948 (13) | Ref | - |
| **Age group (years)** | N=11,495 |  |  |
| <5 | 28/156 (18) | 9.3 (5.2-16.8) | **<0.001** |
| 5-12 | 10/622 (2) | 0.8 (0.4-1.7) | 0.518 |
| 13-18 | 13/669 (2) | 0.9 (0.4-1.7) | 0.683 |
| 19-24 | 25/1,115 (2) | Ref | - |
| 25-39 | 161/4,969 (3) | 1.5 (1.0-2.3) | 0.069 |
| 40-59 | 77/3,156 (2) | 1.0 (0.6-1.6) | 0.937 |
| ≥60 | 68/808 (8) | 3.1 (1.9-5.0) | **<0.001** |
| **Sex** | N=11,404 |  |  |
| Male | 144/5,029 (3) | Ref | - |
| Female | 235/6,375 (4) | 1.3 (1.1-1.6) | **0.016** |
| **Province** | N=11,353 |  |  |
| Eastern Cape | 0/9 (0) | - | - |
| Free State | 9/122 (7) | 1.4 (0.6-3.6) | 0.433 |
| Gauteng | 285/9,507 (3) | 2.0 (1.0-3.7) | **0.037** |
| KwaZulu-Natal | 40/534 (7) | 2.4 (1.2-4.7) | **0.013** |
| Limpopo | 6/257 (2) | 1.7 (0.6-4.8) | 0.330 |
| Mpumalanga | 4/241 (2) | 1.3 (0.4-4.1) | 0.710 |
| North West | 11/274 (4) | 2.9 (1.2-6.9) | **0.019** |
| Northern Cape | 4/108 (4) | 0.8 (0.2-2.4) | 0.638 |

|  |  |  |  |  |
| --- | --- | --- | --- | --- |
|  | Western Cape | 12/301 (4) | Ref | - |
| **Healthcare sector** |  | N=11,495 |  |  |
|  | Public | 159/3,026 (5) | Ref | - |
|  | Private | 223/8,469 (3) | 0.8 (0.6-1.0) | **0.024** |
| **Re-infection<sup>c</sup>** |  | N=11,495 |  |  |
|  | No | 351/10,352 (3) | Ref | - |
|  | Yes | 31/1,143 (3) | 1.1 (0.7-1.6) | 0.660 |

<sup>a</sup> Cases followed-up for hospital admission until 21 December 2021
<sup>b</sup> Admission to hospital between 7 days prior to 21 days after diagnosis (specimen collection date)
<sup>c</sup> Re-infection was defined as an individual with at least one positive SARS-CoV-2 test >90 days prior to the current episode

Among hospitalised individuals diagnosed between 1 October – 30 November 2021, 83.0% (317/382) had accumulated in-hospital outcome by 21 December. After controlling for factors associated with severe COVID-19 disease, the odds of severe disease did not differ between individuals with SGTF infection compared to non-SGTF infections (aOR 0.7, 95% CI 0.3-1.4) (Table 2). The odds of severe disease varied geographically and was higher among individuals aged ≥60 years (aOR 11.5, 95% CI 2.8-47.0), compared to individuals aged 19-24 years.

**Table 2.**
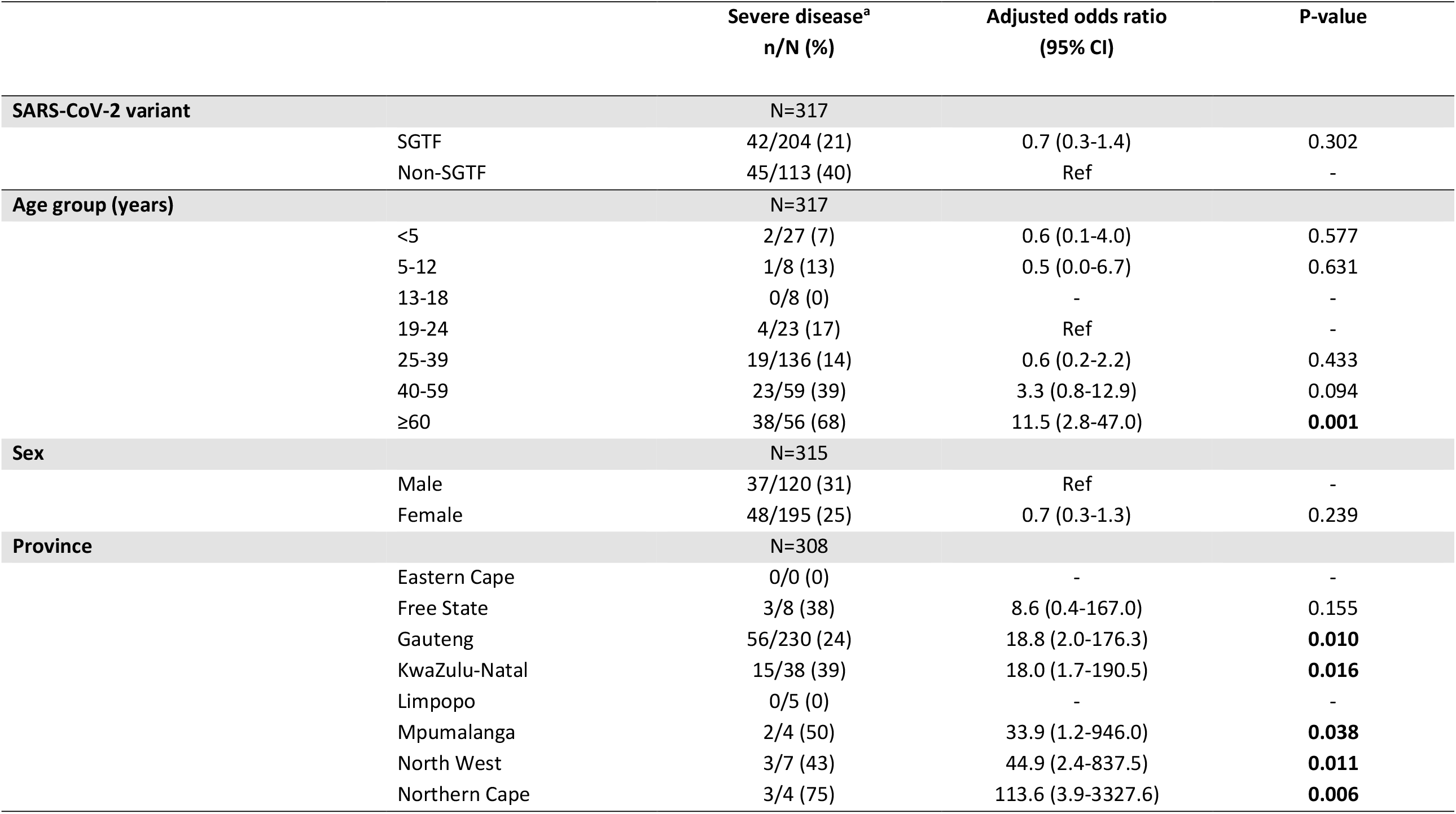
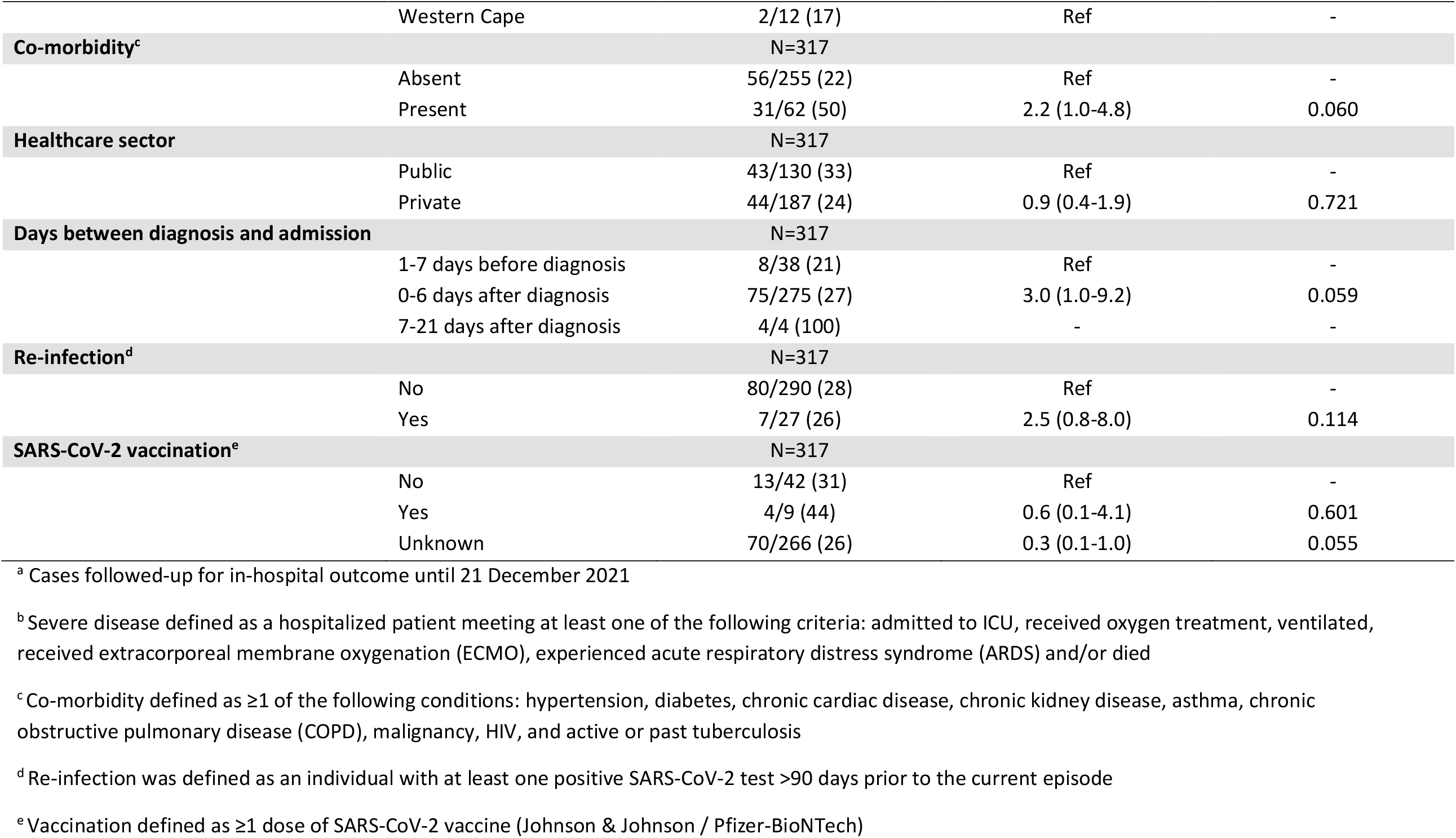
Multivariable logistic regression analysis evaluating the association between S gene target failure (SGTF) infection, compared to non-SGTF infection, and severe disease among hospitalised individuals with known outcome, South Africa, 1 October – 30 November 2021^a^ (N=290)

|  | Severe disease<sup>a</sup><br>n/N (%) | Adjusted odds ratio<br>(95% CI) | P-value |
| --- | --- | --- | --- |
| **SARS-CoV-2 variant** |  |  |  |
|  | N=317 |  |  |
| SGTF | 42/204 (21) | 0.7 (0.3-1.4) | 0.302 |
| Non-SGTF | 45/113 (40) | Ref | - |
| **Age group (years)** |  |  |  |
|  | N=317 |  |  |
| <5 | 2/27 (7) | 0.6 (0.1-4.0) | 0.577 |
| 5-12 | 1/8 (13) | 0.5 (0.0-6.7) | 0.631 |
| 13-18 | 0/8 (0) | - | - |
| 19-24 | 4/23 (17) | Ref | - |
| 25-39 | 19/136 (14) | 0.6 (0.2-2.2) | 0.433 |
| 40-59 | 23/59 (39) | 3.3 (0.8-12.9) | 0.094 |
| ≥60 | 38/56 (68) | 11.5 (2.8-47.0) | **0.001** |
| **Sex** |  |  |  |
|  | N=315 |  |  |
| Male | 37/120 (31) | Ref | - |
| Female | 48/195 (25) | 0.7 (0.3-1.3) | 0.239 |
| **Province** |  |  |  |
|  | N=308 |  |  |
| Eastern Cape | 0/0 (0) | - | - |
| Free State | 3/8 (38) | 8.6 (0.4-167.0) | 0.155 |
| Gauteng | 56/230 (24) | 18.8 (2.0-176.3) | **0.010** |
| KwaZulu-Natal | 15/38 (39) | 18.0 (1.7-190.5) | **0.016** |
| Limpopo | 0/5 (0) | - | - |
| Mpumalanga | 2/4 (50) | 33.9 (1.2-946.0) | **0.038** |
| North West | 3/7 (43) | 44.9 (2.4-837.5) | **0.011** |
| Northern Cape | 3/4 (75) | 113.6 (3.9-3327.6) | **0.006** |

|  |  |  |  |  |
| --- | --- | --- | --- | --- |
|  | Western Cape | 2/12 (17) | Ref | - |
| **Co-morbidity<sup>c</sup>** |  | N=317 |  |  |
|  | Absent | 56/255 (22) | Ref | - |
|  | Present | 31/62 (50) | 2.2 (1.0-4.8) | 0.060 |
| **Healthcare sector** |  | N=317 |  |  |
|  | Public | 43/130 (33) | Ref | - |
|  | Private | 44/187 (24) | 0.9 (0.4-1.9) | 0.721 |
| **Days between diagnosis and admission** |  | N=317 |  |  |
|  | 1-7 days before diagnosis | 8/38 (21) | Ref | - |
|  | 0-6 days after diagnosis | 75/275 (27) | 3.0 (1.0-9.2) | 0.059 |
|  | 7-21 days after diagnosis | 4/4 (100) | - | - |
| **Re-infection<sup>d</sup>** |  | N=317 |  |  |
|  | No | 80/290 (28) | Ref | - |
|  | Yes | 7/27 (26) | 2.5 (0.8-8.0) | 0.114 |
| **SARS-CoV-2 vaccination<sup>e</sup>** |  | N=317 |  |  |
|  | No | 13/42 (31) | Ref | - |
|  | Yes | 4/9 (44) | 0.6 (0.1-4.1) | 0.601 |
|  | Unknown | 70/266 (26) | 0.3 (0.1-1.0) | 0.055 |

<sup>a</sup> Cases followed-up for in-hospital outcome until 21 December 2021
<sup>b</sup> Severe disease defined as a hospitalized patient meeting at least one of the following criteria: admitted to ICU, received oxygen treatment, ventilated, received extracorporeal membrane oxygenation (ECMO), experienced acute respiratory distress syndrome (ARDS) and/or died
<sup>c</sup> Co-morbidity defined as $\geq 1$ of the following conditions: hypertension, diabetes, chronic cardiac disease, chronic kidney disease, asthma, chronic obstructive pulmonary disease (COPD), malignancy, HIV, and active or past tuberculosis
<sup>d</sup> Re-infection was defined as an individual with at least one positive SARS-CoV-2 test >90 days prior to the current episode
<sup>e</sup> Vaccination defined as $\geq 1$ dose of SARS-CoV-2 vaccine (Johnson & Johnson / Pfizer-BioNTech)

### Delta vs. SGTF infection analysis

From 31 March 2020 through 6 December 2021, there were 1,734 hospitalised patients for whom variant information was available either by genome sequencing (non-variant, Alpha, Beta, Delta), or TaqPath PCR (SGTF, as a proxy for Omicron from 1 October through 6 December). Of these, 792 (46%) were infected with the Delta variant from 1 April 2021 (week 13) to 9 November 2021 (week 45), and 212 (12%) with SGTF from 1 October 2021 (week 42) to 6 December 2021 (week 48) (Figure 2).

**Figure 2.**
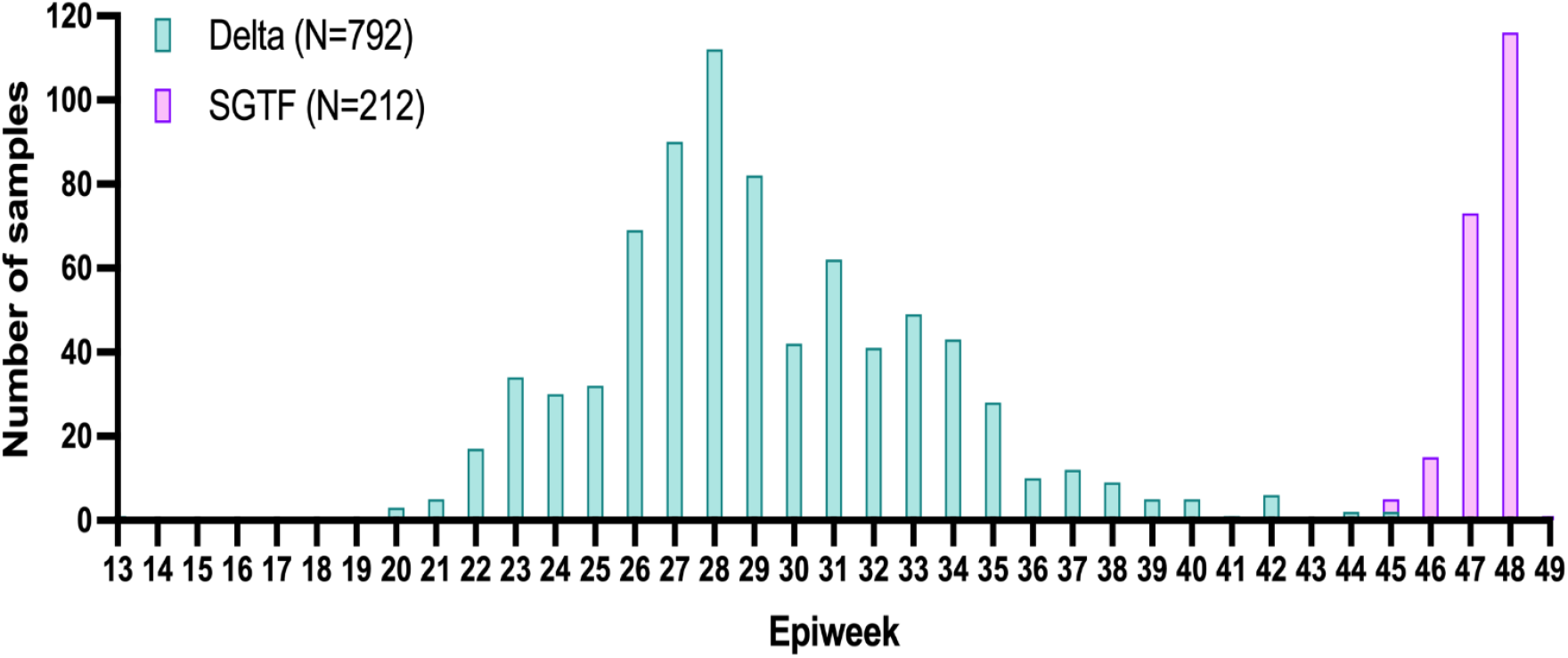
Number of SARS-CoV-2 Delta variant and S Gene Target Failure (SGTF) specimens among hospitalized COVID-19 cases with known outcome by epidemiological week and variant type, South Africa DATCOV-Gen, 3 April (week 13) - 6 December (week 48)

Among hospitalised individuals, 90.8% (1,037/1,142) had accumulated in-hospital outcome by 21 December 2021. After controlling for factors associated with severe disease, SGTF infections diagnosed between 1 October and 30 November 2021 compared to Delta infections diagnosed between April and November 2021 had a lower odds of severe disease (aOR 0.3, 95% CI 0.2-0.5) (Table 3). In addition to geographical differences, other factors identified as associated with an increased odds of severe disease included older age [40-59 years (aOR 2.2, 95% CI 1.0-5.0) and ≥60 years (aOR 3.8, 95% CI 1.7-8.6), compared to 19-24 years] and having a co-morbid condition (aOR 2.8, 95% CI 2.0-4.0). Individuals aged 13-18 years (aOR 0.2, 95% CI 0.0-0.8, compared to 19-24 years) and females (aOR 0.7, 95% CI 0.5-1.0) had a lower odds of severe disease.

**Table 3.**
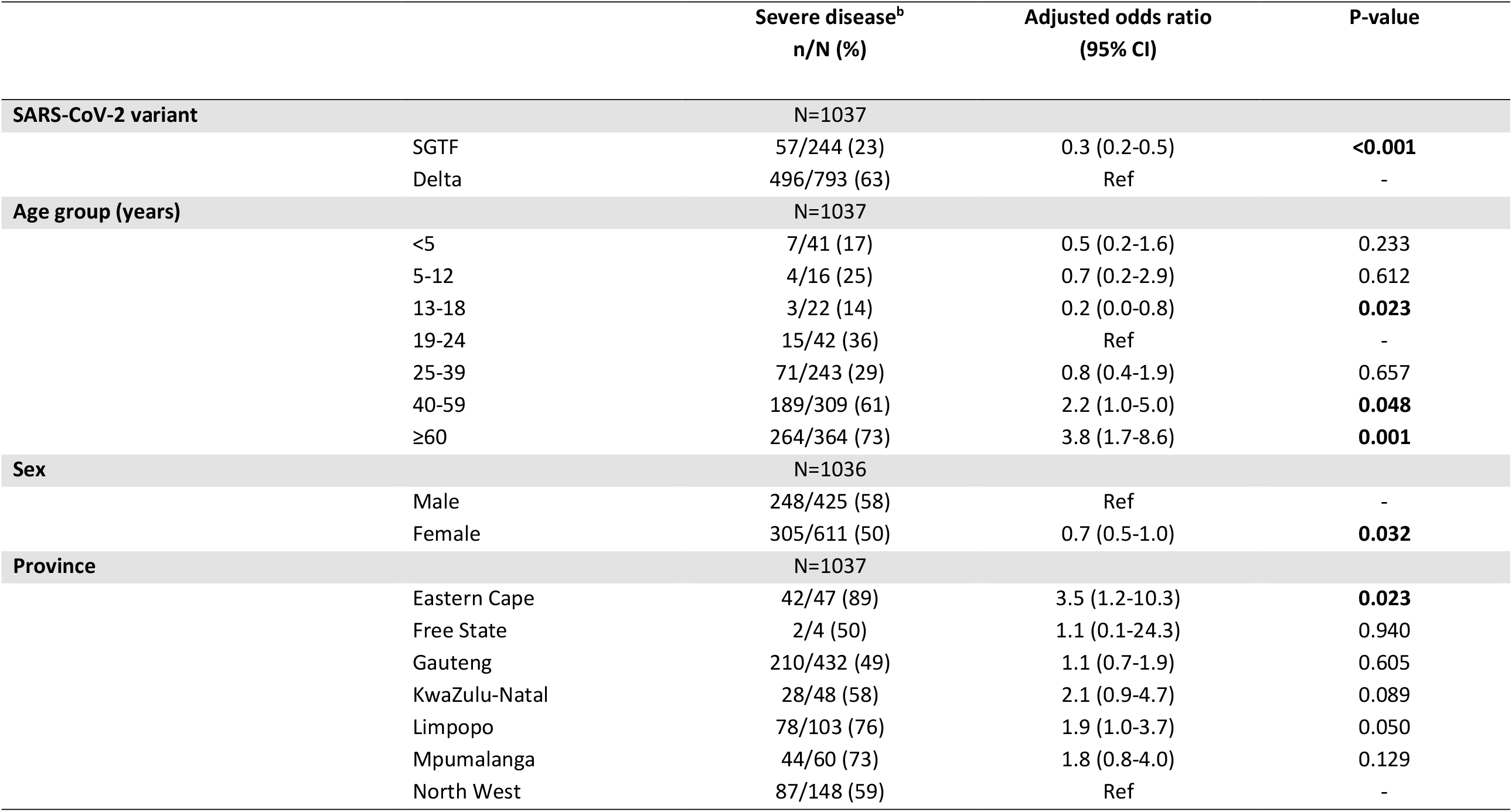
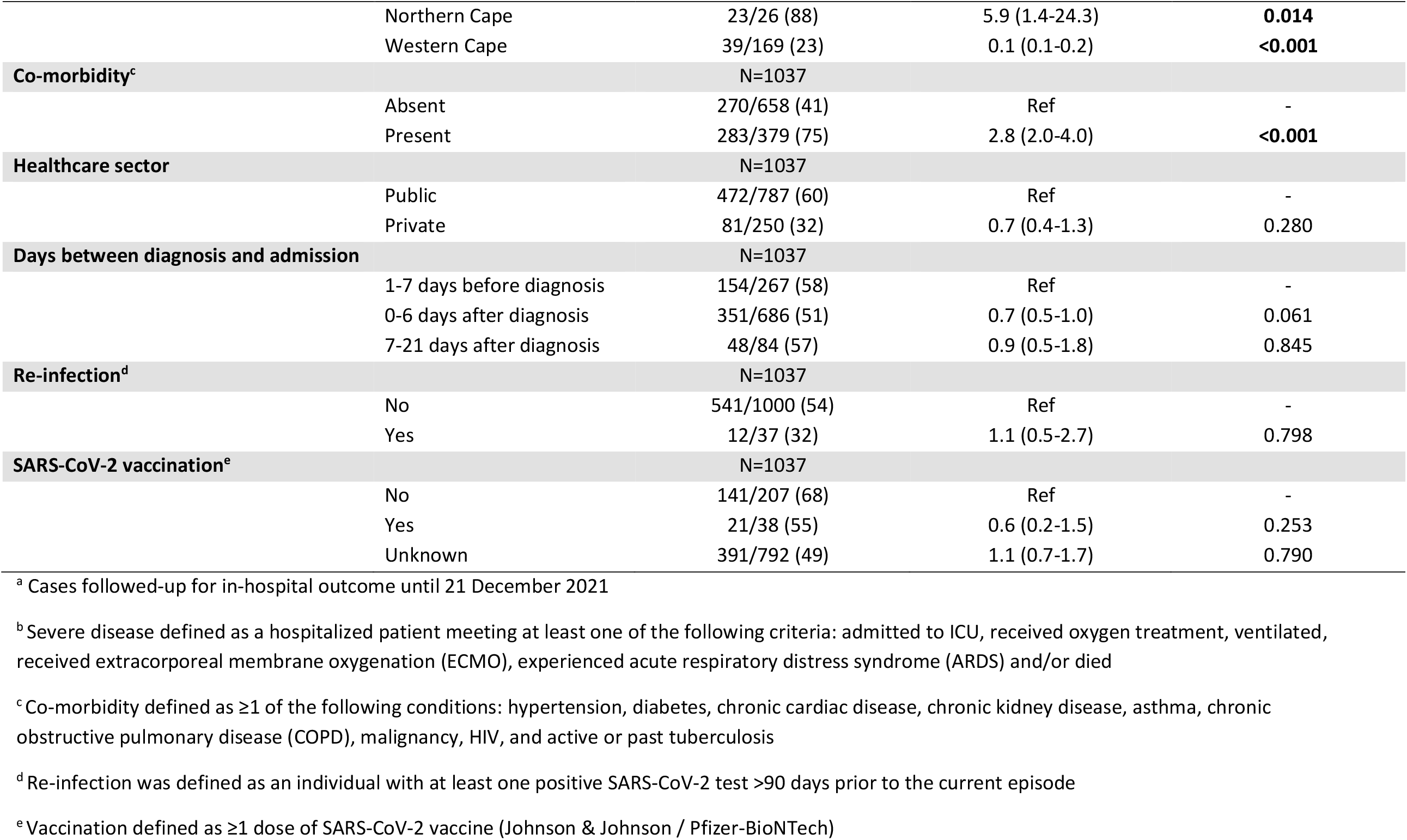
Multivariable logistic regression analysis evaluating the association between S gene target failure (SGTF) infection during 1 October – 30 November 2021, compared to Delta variant infection during April – November 2021, and severe disease among hospitalized individuals with known outcome, South Africa^a^ (N=1036)

|  | Severe disease<sup>b</sup><br>n/N (%) | Adjusted odds ratio<br>(95% CI) | P-value |
| --- | --- | --- | --- |
| **SARS-CoV-2 variant** N=1037 |  |  |  |
| SGTF | 57/244 (23) | 0.3 (0.2-0.5) | **<0.001** |
| Delta | 496/793 (63) | Ref | - |
| **Age group (years)** N=1037 |  |  |  |
| <5 | 7/41 (17) | 0.5 (0.2-1.6) | 0.233 |
| 5-12 | 4/16 (25) | 0.7 (0.2-2.9) | 0.612 |
| 13-18 | 3/22 (14) | 0.2 (0.0-0.8) | **0.023** |
| 19-24 | 15/42 (36) | Ref | - |
| 25-39 | 71/243 (29) | 0.8 (0.4-1.9) | 0.657 |
| 40-59 | 189/309 (61) | 2.2 (1.0-5.0) | **0.048** |
| ≥60 | 264/364 (73) | 3.8 (1.7-8.6) | **0.001** |
| **Sex** N=1036 |  |  |  |
| Male | 248/425 (58) | Ref | - |
| Female | 305/611 (50) | 0.7 (0.5-1.0) | **0.032** |
| **Province** N=1037 |  |  |  |
| Eastern Cape | 42/47 (89) | 3.5 (1.2-10.3) | **0.023** |
| Free State | 2/4 (50) | 1.1 (0.1-24.3) | 0.940 |
| Gauteng | 210/432 (49) | 1.1 (0.7-1.9) | 0.605 |
| KwaZulu-Natal | 28/48 (58) | 2.1 (0.9-4.7) | 0.089 |
| Limpopo | 78/103 (76) | 1.9 (1.0-3.7) | 0.050 |
| Mpumalanga | 44/60 (73) | 1.8 (0.8-4.0) | 0.129 |
| North West | 87/148 (59) | Ref | - |

|  |  |  |  |  |
| --- | --- | --- | --- | --- |
|  | Northern Cape | 23/26 (88) | 5.9 (1.4-24.3) | **0.014** |
|  | Western Cape | 39/169 (23) | 0.1 (0.1-0.2) | **<0.001** |
| **Co-morbidity<sup>c</sup>** |  | N=1037 |  |  |
|  | Absent | 270/658 (41) | Ref | - |
|  | Present | 283/379 (75) | 2.8 (2.0-4.0) | **<0.001** |
| **Healthcare sector** |  | N=1037 |  |  |
|  | Public | 472/787 (60) | Ref | - |
|  | Private | 81/250 (32) | 0.7 (0.4-1.3) | 0.280 |
| **Days between diagnosis and admission** |  | N=1037 |  |  |
|  | 1-7 days before diagnosis | 154/267 (58) | Ref | - |
|  | 0-6 days after diagnosis | 351/686 (51) | 0.7 (0.5-1.0) | 0.061 |
|  | 7-21 days after diagnosis | 48/84 (57) | 0.9 (0.5-1.8) | 0.845 |
| **Re-infection<sup>d</sup>** |  | N=1037 |  |  |
|  | No | 541/1000 (54) | Ref | - |
|  | Yes | 12/37 (32) | 1.1 (0.5-2.7) | 0.798 |
| **SARS-CoV-2 vaccination<sup>e</sup>** |  | N=1037 |  |  |
|  | No | 141/207 (68) | Ref | - |
|  | Yes | 21/38 (55) | 0.6 (0.2-1.5) | 0.253 |
|  | Unknown | 391/792 (49) | 1.1 (0.7-1.7) | 0.790 |

<sup>a</sup> Cases followed-up for in-hospital outcome until 21 December 2021
<sup>b</sup> Severe disease defined as a hospitalized patient meeting at least one of the following criteria: admitted to ICU, received oxygen treatment, ventilated, received extracorporeal membrane oxygenation (ECMO), experienced acute respiratory distress syndrome (ARDS) and/or died
<sup>c</sup> Co-morbidity defined as ≥1 of the following conditions: hypertension, diabetes, chronic cardiac disease, chronic kidney disease, asthma, chronic obstructive pulmonary disease (COPD), malignancy, HIV, and active or past tuberculosis
<sup>d</sup> Re-infection was defined as an individual with at least one positive SARS-CoV-2 test >90 days prior to the current episode
<sup>e</sup> Vaccination defined as ≥1 dose of SARS-CoV-2 vaccine (Johnson & Johnson / Pfizer-BioNTech)

## DISCUSSION

In November 2021, the Omicron variant emerged in South Africa, concurrent with a rapid increase in the number of reported COVID-19 cases^1^ leading to the fourth epidemic wave. The identification of the new variant caused concern due to the large number of mutations present throughout the genome. In particular, mutations that are known to be associated with immune escape and increased infectivity were identified in the spike protein. While a number of studies are underway to rapidly assess phenotypic behaviour and immune escape, it is also critical to understand the clinical severity of infections caused by Omicron. We evaluated the severity of SGTF infections (as a proxy for the Omicron variant) by comparing to (i) non-SGTF infections during the same period of time, and (ii) Delta variant infections, which dominated the third wave. When compared to non-SGTF infections, we found that SGTF infections had an 80% lower odds being admitted to hospital, but did not differ in the risk of severe disease among hospitalised individuals. When compared to Delta infections, SGTF infections were associated with a 70% lower odds of severe disease. In addition, we found that early Omicron wave infections had a significantly lower mean PCR Ct-value compared to early Delta wave infections, which may reflect higher viral loads in Omicron infected individuals.

Our findings correlate with DATCOV surveillance hospitalisation data showing that, among patients who had a known hospital outcome, 32% of COVID-19 admissions during the early fourth wave were severe compared to 65% during the early third wave^10^. By November 2021, a high proportion of the South African population had some level of SARS-CoV-2 immunity, as a result of previous natural infection and/or vaccination. It is estimated that after the third SARS-CoV-2 wave 60%-70% of individuals in South Africa had evidence of previous SARS-CoV-2 infection^11^. By 9 December 2021, 65% of individuals aged ≥60 years, 61% aged 50-59 years, 48% aged 35-49 and 29% aged 18-34 years had received ≥1 dose of SARS-CoV-2 vaccine, and 58% of individuals aged ≥60 years, 55% aged 50-59 years, 43% aged 35-49 and 24% aged 18-34 years were fully vaccinated against SARS-CoV-2 (one dose of Johnson & Johnson or two doses of Pfizer-BioNTech)^12^. It is difficult to disentangle the relative contribution of high levels of previous population immunity versus intrinsic lower virulence to the observed lower disease severity. Our finding of no difference in severity in SGTF compared to non-SGTF infected individuals in the same time period, and the lower risk of severity in SGTF compared to earlier Delta infected individuals, suggests that this reduced severity may be in part a result of high levels of population immunity (due to natural infection and/or vaccination). Incomplete vaccination data, and the fact that the majority of re-infections were likely not detected, resulted in incomplete adjustment for the effect of prior immunity in our analyses.

This study has a number of limitations. Firstly, SGTF infections were only identifiable using the TaqPath PCR and only for specimens with high viral loads (Ct≤30) and therefore the number of SGTF infections is underestimated and biased towards geographic regions where this assay was more commonly used. Secondly, SGTF identified on PCR was used as a proxy for Omicron variant detection. However, SGTF may also identify the Alpha variant and occur sporadically in other variants and therefore some infections may have been misclassified as Omicron. However, we only used SGTF as a proxy for Omicron after 1 October 2021 and genome data generated by NGS-SA has not identified the Alpha variant in South Africa since August 2021 and at its peak Alpha variant was only detected in 6% of samples in May 2021 ^13^. In addition, Omicron has recently been classified into three sub-lineages, one (BA.2) of which does not contain the Δ69-70 deletion and which therefore will not be identifiable by the SGTF. Genome data from November 2021, showed that of 881 Omicron sequences, 872 (99.0%) were BA.1, 1 (0.1%) was BA.2 and 8 (0.9%) were BA.3^13^. Ongoing sequencing will enable definitive classification of the Omicron variant for future severity analyses. Thirdly, our analysis was conducted in the early phase of the fourth wave after the emergence of Omicron when numbers were small, patients with milder symptoms were more likely to be admitted, and there may be a lag in hospitalisations and severe outcomes caused by this new variant. We accounted for this by only including hospitalised patients with known outcomes, censoring cases to ensure at least three weeks of follow up, and adjusting for time interval between diagnosis and hospitalisation in the severity multivariable models. The inclusion of individuals only with accumulated in-hospital outcomes may have biased SGTF towards shorter hospital stay, hence this result should be interpreted with caution. Fourthly, we compared cases through the full Delta wave to cases in the ascending phase of the Omicron wave, this could bias comparisons if case characteristics differ in the ascending and descending wave phases. However, data from the DATCOV programme suggest that the proportion of severe cases does not vary substantially through the different wave phases^10^. Lastly, we had limited vaccination information only for hospitalised cases and that was based on self-report.

Despite the Omicron variant being only recently detected, there has been a rapid increase in the prevalence of SGTF infections across all provinces during November 2021, with almost complete replacement of Delta over a period of four weeks. Early analyses indicate a reduced risk of hospital admission among SGTF-infected individuals when compared to non-SGTF infected individuals diagnosed during October – November 2021, and a reduced risk of severe disease among SGTF-infected individuals when compared to earlier Delta-infected individuals. The high population immunity (due to prior infection and/or vaccination) may be protective against severe disease in our population. These are early data and findings may change as the epidemic progresses, and with additional follow-up time for the more recently diagnosed SGTF-infected individuals.

## Data Availability

All data produced in the present study are available upon reasonable request to the authors

## ACKNOWLEDGEMENTS

We acknowledge additional NGS-SA members: Adriano Mendes, Allison Glass, Amy Strydom, Andiswa Simane, Annabel Enoch, Arash Iranzadeh, Ashlyn Davis, Bulelani Manene, Carolyn Williamson, Cheryl Cohen, Deelan Doolabh, Derek Tshiabuila, Diana Hardie, Dominique Goadhals, Eduan Wilkinson, Gert van Zyl, Houriiyah Tegally, James Emmanuel San, Jennifer Giandhari, Kamela Mahlakwane, Kathleen Subramoney, Kruger Marais, Linda de Gouveia, Louizer Ingasia, Marietjie Venter, Martin Nyaga, Michaela Davids, Nokuzola Ndebele, Noluthando Duma, Oluwakemi Laguda-Akingba, Rageema Joseph, Richard Lessells, Sibongile Walaza, Simnikiwe Mayaphi, Stephen Korsman, Sureshnee Pillay, Susan Engelbrecht, Tania Stander, Terry Marshall, Tulio de Oliveira, Upasana Ramphal, Wolfgang Preiser, Yajna Ramphal, Yeshnee Naidoo, Zinhle Makatini.

We would like to acknowledge the teams within the National Institute for Communicable Diseases Centre for Respiratory Diseases and Meningitis (Linda de Gouveia, Thabo Mohale, Boitshoko Mahlangu, Noxolo Ntuli, Anele Mnguni, Gerald Motsatsi, Malusi Ndlovu, Noluthando Duma), and Sequencing Core Facility (Annie Chan, Morne du Plessis). We thank Nevashan Govender, Genevie Ntshoe, Andronica Moipone Shonhiwa, Darren Muganhiri, Itumeleng Matiea, Eva Mathatha, Fhatuwani Gavhi, Teresa Mashudu Lamola, Matimba Makhubele, Chidozie Iwu, Simbulele Mdleleni, Masingita Makhubela from the national SARS-CoV-2 NICD surveillance team; Fazil Mckenna, Trevor Graham Bell, Ndivhuwo Munava, Stanford Kwenda, Muzammil Raza Bano and Jimmy Khosa from NICD IT for NMCSS case data. We thank all laboratories for submitting specimens for sequencing, and all public laboratories and Lancet Laboratories for Thermo Fisher TaqPath PCR data. We thank all hospitals and healthcare workers submitting data through the DATCOV surveillance programme. We are grateful to Cecile Viboud and Kaiyuan Sun of the Fogarty International Center, National Institutes of Health, USA for their input on the analysis.

This study was funded by the South African Medical Research Council with funds received from the National Department of Health. Sequencing activities for NICD are supported by a conditional grant from the South African National Department of Health as part of the emergency COVID-19 response; a cooperative agreement between the National Institute for Communicable Diseases of the National Health Laboratory Service and the United States Centers for Disease Control and Prevention (grant number 5 U01IP001048-05-00); the African Society of Laboratory Medicine (ASLM) and Africa Centers for Disease Control and Prevention through a sub-award from the Bill and Melinda Gates Foundation grant number INV-018978; the UK Foreign, Commonwealth and Development Office and Wellcome (Grant no 221003/Z/20/Z); the South African Medical Research Council (Reference number SHIPNCD 76756); and the UK Department of Health and Social Care and managed by the Fleming Fund and performed under the auspices of the SEQAFRICA project. The Fleming Fund is a £265 million UK aid programme supporting up to 24 low- and middle-income countries (LMICs) generate, share and use data on antimicrobial resistance (AMR) and works in partnership with Mott MacDonald, the Management Agent for the Country and Regional Grants and Fellowship Programme. This research was also supported by The Coronavirus Aid, Relief, and Economic Security Act (CARES ACT) through the Centers for Disease Control and Prevention (CDC) and the COVID International Task Force (ITF) funds through the CDC under the terms of a subcontract with the African Field Epidemiology Network (AFENET) AF-NICD-001/2021. Screening for SGTF at UCT was supported by the Wellcome Centre for Infectious Diseases Research in Africa (CIDRI-Africa), which is supported by core funding from the Wellcome Trust (203135/Z/16/Z and 222754).

The findings and conclusions in this report are those of the author(s) and do not necessarily represent the official position of the funding agencies.

